# Peritumoral *SULF1* Transcript Signal Is Associated with Disease-Specific Survival in Oral Squamous Cell Carcinoma

**DOI:** 10.64898/2026.10.09.26364914

**Authors:** Gregory Furletti, Trystan Conrad Guerrero, Edward C. Schrom, Bruce Davidson, Bhaskar Kallakury, Radoslav Goldman

**Author notes:** Corresponding author: Radoslav Goldman, Department of Oncology, Georgetown University Medical Center, 3800 Reservoir Road NW, Washington, DC 20057, USA.

## Abstract

The prognostic relevance of peritumoral *SULF1* transcript signal in oral squamous cell carcinoma (OSCC) is unclear. We assembled a retrospective cohort of 133 patients with 218 primary-site tissue-microarray cores; 131 patients had evaluable boundary features. Multiplex fluorescence imaging combined RNAscope *SULF1*, CK-pan, and DAPI. Tumor-stroma boundaries were annotated with the *SULF1* channel hidden. We screened 50 exported boundary features across cumulative stromal bands of 35-300 μm against recurrence-free survival (RFS), overall survival (OS), and disease-specific survival (DSS), with false-discovery-rate correction. The strongest DSS association was signal SD within 150 μm (n = 107; 23 disease-specific deaths; HR = 2.20 per SD; 95% CI, 1.64-2.94; q = 2.1 × 10⁻⁶). In 50 repeated 5-fold cross-validations with feature selection repeated within training folds, adding a boundary feature to AJCC stage increased mean held-out DSS c-index from 0.613 to 0.754 (ΔC = +0.141; 2.5th-97.5th percentiles across repeats, +0.105 to +0.192). These percentiles describe split variability, not confidence intervals. The 150-μm SD feature was selected in 39.2% of DSS training folds. The DSS association persisted after adjustment for mean signal, although the measurements were strongly correlated. Peritumoral *SULF1* signal dispersion was associated with outcome beyond an ordinal AJCC-stage model; independent validation is required.

## Introduction

GLOBOCAN 2022 estimated approximately 390,000 new cases of lip and oral cavity cancer worldwide.^1^ Outcomes remain variable for early-stage oral squamous cell carcinoma (OSCC). In a series of 216 cT1-T2N0 oral-tongue OSCCs treated with surgery, five-year recurrence-free survival was 70%.^2^ American Joint Committee on Cancer (AJCC) stage and depth of invasion inform risk assessment,^3–6^ and tumor budding and worst pattern of invasion provide additional prognostic information in early oral-tongue SCC.^7^ These clinicopathologic features do not fully explain variation in outcome.^2,7^

SULF-1 is an extracellular heparan sulfate 6-O-endosulfatase that modifies the sulfation of cell-surface and extracellular-matrix heparan sulfate proteoglycans (HSPGs).^8–10^ HSPGs bind growth factors, cytokines, chemokines, and morphogens and regulate their extracellular distribution and receptor signaling.^11–12^ In a quail model, QSulf1-mediated 6-O-desulfation reduced Wnt sequestration by HSPGs and promoted Wnt receptor signaling.^13^ We use *SULF1* for the gene or transcript and SULF-1 for the protein. Reanalysis of the Puram et al. head and neck squamous cell carcinoma (HNSCC) single-cell dataset showed that *SULF1* expression was concentrated in fibroblasts, whereas *SULF2* was expressed mainly by tumor cells.^8,14^ RNAscope localized *SULF1* transcript mainly to cytokeratin-negative stroma.^8^ CRISPR/Cas9 knockout of *SULF1* in primary HNSCC cancer-associated fibroblasts (CAFs) reduced CAF-supported cancer cell migration and invasion in vitro.^15^ In colorectal cancer, *SULF1*-expressing CAFs promote VEGFA-dependent extracellular matrix remodeling and angiogenesis, and high stromal *SULF1* expression is associated with poor prognosis.^16^ CAFs comprise heterogeneous populations with roles in matrix remodeling, soluble-factor signaling, and immune regulation.^17–19^

Yang et al. localized *SULF1* transcript mainly to the stroma in OSCC and compared node-positive with node-negative tumors, but did not analyze signal as a function of distance from the tumor-stroma boundary.^8^ In their analysis of The Cancer Genome Atlas HNSCC cohort, high *SULF1* mRNA was associated with shorter progression-free interval in early-stage disease after adjustment for *SULF2* and clinical covariates; no association was detected in advanced-stage disease.^8^

We examined whether peritumoral *SULF1* transcript signal varies with distance from the tumor-stroma boundary and whether its intensity and dispersion are associated with recurrence-free survival (RFS), overall survival (OS), and disease-specific survival (DSS) in OSCC. We also tested whether boundary measurements remain associated with outcome after adjustment for AJCC stage and mean stromal signal.

## Materials and Methods

### Study Cohort and Tissue Microarrays

We reviewed 183 clinical records. Eighteen records were excluded because their tissue-processing results were unavailable, leaving 165. Fifteen lacked qualifying linked primary-site tissue and 11 contributed only lymph-node or metastatic cores, leaving 139 records. Shared case and specimen identifiers linked these records to 138 distinct patients with 224 primary-site cores. We excluded one patient with salivary carcinoma (two cores), three patients with in-situ SCC (three cores), and one additional patient recorded as Stage CIS (one core). The eligible invasive-OSCC cohort therefore comprised 133 patients and 218 primary-site cores. Boundary measurements were available for 215 cores from 131 patients. The 46 lymph-node or metastatic cores were excluded from analysis of the primary-site cohort. Core measurements were averaged within each patient, including patients represented by multiple specimens. The patient was the unit of inference.

Tissue microarrays (TMAs) were constructed from formalin-fixed paraffin-embedded (FFPE) surgical resection specimens. Six TMAs were sampled with 1 to 4 cores per patient (mean, 1.64 primary-site cores per eligible patient) depending on available tissue. Tumors were staged according to the American Joint Committee on Cancer (AJCC) 8th edition staging system.^3^

Clinical variables collected included age at diagnosis, sex, tobacco use history, primary site, pathologic T-stage, N-stage, overall AJCC stage, histologic grade, depth of invasion, and margin status. Follow-up data included dates and events for recurrence, death, and cause of death. The study was approved by the institutional review board (IRB 1992-48). Informed consent was obtained from all participants. We followed the REMARK guidelines^20^ for reporting tumor marker prognostic studies throughout the design, analysis, and reporting of this work.

The retained specimens were accessioned from 1995 through 2020. Analyses used the follow-up durations recorded for each patient; the latest recorded follow-up date among eligible patients was March 28, 2022. We identified primary-site tumor cores using the specimen and tissue labels recorded for each core. A tissue label from another specimen belonging to the same patient was not used to classify the imaged core.

### Multiplex Fluorescence Staining and RNAscope

Sequential FFPE TMA sections (4 μm) underwent multiplex fluorescence imaging that combined pan-cytokeratin (CK-pan; Opal 520) with RNAscope detection of *SULF1* transcript (Advanced Cell Diagnostics probe 403581-C3; C3/Opal 690) and DAPI nuclear counterstaining. Slides were processed according to the manufacturers’ protocols. Positive and negative controls were included in the fluorescence assay and assessed by visual review.

Slides were scanned on a Vectra Polaris automated quantitative pathology imaging system (Akoya Biosciences) using Fusion v1.0.5, 14-bit acquisition, 20× magnification, and 0.50 μm/pixel resolution. Whole-slide images were exported to QuPath. Manually annotated tumor regions defined the tumor and stromal compartments. All six TMA slides were scanned in one imaging batch under the same acquisition settings but were not stained in a single batch.

Boundary features were calculated from exported per-cell signal without post hoc per-TMA normalization; continuous predictors were z-score standardized within each endpoint-specific analytic sample for regression. Potential staining-run confounding was assessed in sensitivity Cox models with baseline hazards stratified by TMA number.

### Image Analysis, Cell Segmentation, and Boundary Annotation

Cell detection and segmentation were performed in QuPath^21^ v0.6.0 using the DAPI channel and QuPath’s built-in watershed-based cell detection; no StarDist model was used. Cell detection used a requested pixel size of 0.5 μm, background radius of 8 μm with opening by reconstruction enabled, median filter radius of 0 μm, sigma of 1.5 μm, nuclear area range of 10-400 μm², intensity threshold of 25, split-by-shape enabled, cell expansion of 5 μm, inclusion of the nucleus within the cell object, boundary smoothing enabled, and measurement generation enabled. Cell centroids, mean DAPI and CK-pan signals, and mean per-cell *SULF1* transcript signal were exported for each TMA core. Tumor and stromal compartment labels were not assigned by a global CK-pan intensity threshold. Instead, tumor membership was defined by manually annotated tumor polygons: cells inside clipped tumor annotations were labeled tumor, and cells outside these polygons were labeled stroma for boundary-distance analyses and for SPACE (Spatial Patterning Analysis of Cellular Ensembles).

A diagnostic hematoxylin and eosin (H&E)-stained section was reviewed for each case to confirm invasive OSCC and identify the region used for coring; depth of invasion was measured on whole-slide H&E according to AJCC 8th-edition criteria. Tumor boundaries were drawn in QuPath with the *SULF1* channel hidden, using matched H&E, CK-pan, and DAPI for orientation. An independent review by a blinded pathologist qualitatively confirmed the annotations before feature extraction. No numeric inter-reader agreement statistic was calculated.

After boundary annotations were finalized, cell-level marker intensities were extracted for all channels. A core viability assessment was performed to identify and exclude non-viable cores (those with tissue folding, loss, poor staining quality, or insufficient tumor content). The viability mapping was recorded before any analysis, and non-viable cores were excluded from all downstream computations (cohort derivation is given above).

### Boundary Distance Calculation

Tumor boundary annotations were exported from QuPath as GeoJSON polygons and converted to the cell-centroid coordinate system. For each cell, we computed the signed Euclidean distance from the nuclear centroid to the nearest boundary point. Positive values indicate cells on the stromal side of the boundary, negative values indicate cells within the tumor compartment, and zero represents the interface. Inside/outside classification used a ray-casting point-in-polygon test, and nearest-boundary queries were accelerated with scipy.spatial.cKDTree.

For analytic boundary-band features, cells were assigned to five concentric stromal-side distance thresholds: 35, 70, 105, 150, and 300 μm from the tumor-stroma interface. Each band included all stromal cells from 0 up to the band threshold (cumulative bands, not annular rings). These distances sampled the immediate peritumoral zone through the more distant stromal compartment. For the signed-distance visualization, per-core mean signal in 10-μm bins was averaged within each patient and then across contributing patients. The number of patients contributing to each bin could vary with available tissue. Cell counts in this profile were pooled across cores and were not normalized to sampled tissue area.

### Boundary Band Features

For each core and cumulative stromal band, we calculated *SULF1* signal mean, median, sample SD, 95th percentile (p95), 99th percentile (p99), and mean signal among cells at or above the 90th percentile. Positivity was defined by a two-component Gaussian mixture (GMM) fitted separately within each core-band combination, using the midpoint between component means as the threshold. Positivity required at least 10 cells and a nonconstant signal distribution. The positive-cell fraction and its percentage encoding were both exported. The variable named positive area fraction in the analysis files is a cell fraction; it does not measure stained tissue area. We also recorded cell count and the ordinary least-squares slope of per-cell signal against distance within each band. Quantiles used linear interpolation; no additional minimum cell count was imposed for p99.

Five bands and 10 exported fields per band produced 50 boundary columns. Because positive-cell fraction and percentage encode the same measurement, these represent 45 distinct quantities. Both encodings were retained in the exploratory screen. Patient-level features were the arithmetic mean across available core-level measurements. The 35-μm bands contained a median of 1,557 cells per evaluable core (IQR, 798-2,550; range, 20-6,382); seven cores contained fewer than 100 cells. SD measures dispersion and p99 measures upper-tail intensity; neither is independent of signal scale. Cox models used one row per patient, Efron handling of tied events, and model-based standard errors. Descriptive tumor-stroma comparisons used each patient’s core-averaged mean signal in each compartment; the stromal-to-tumor ratio was calculated from these patient-level means. We summarized the per-patient band slopes and GMM-defined positive-cell fractions by their median and interquartile range. These descriptive analyses used the 131 patients with evaluable boundary measurements. Because GMM thresholds were fitted separately for each core and band, positive-cell fractions did not use a common positivity threshold across compartments or distances.

### Exploratory cis mutual information

We quantified spatial co-organization of *SULF1* transcript signal and tumor or stromal labels using cis mutual information (cisMI), a census-derived spatial-patterning statistic.^22^ All valid exported cells provided neighborhood context; cells within 120 μm on either side of the interface were eligible neighborhood centers. Centers were sampled separately at each radius to target five-fold mean coverage of this boundary domain. A core-radius combination required at least 39 neighborhoods, sufficient for two SPACE bins at ensemble depth 2. For each combination, 999 label-shuffle resamples generated a null mutual-information distribution. cisMI was the observed mutual information minus the mean of this null distribution. We divided cisMI by log2(B), where B was the effective SPACE bin count, and averaged these values across viable primary-site cores within each patient.

Five linked variable families were evaluated at 35, 70, 105, 150, 225, and 300 μm: tumor-linked *SULF1*, stromal-linked *SULF1*, combined tumor- and stromal-linked *SULF1*, tumor-with-tumor-linked *SULF1*, and stromal-with-stromal-linked *SULF1*. Here, tumor-with-tumor-linked cisMI measures the joint spatial pattern of the proportion of cells labeled as tumor and the mean *SULF1* signal of those tumor cells across sampled neighborhoods. These combinations yielded 30 candidate cisMI features. Measurements were available for 215 of the 218 retained primary-site cores; availability varied by radius, and only 29 retained primary-site cores had cisMI measurements at 300 μm. The lead cisMI feature was selected by the lowest univariate P value among candidates with an endpoint-specific sample at least 80% as large as the best-supported candidate. Positive cisMI indicates greater joint spatial organization than the label-shuffled null. cisMI was included in the exploratory screening family but not in the repeated boundary-feature cross-validation.

### Statistical Analysis

All survival analyses used three endpoints: RFS, OS, and DSS, measured in years from diagnosis. OS ended at death from any cause or last verified follow-up for living patients. DSS used the same interval, with DOD (dead of disease) coded as an event; DOC (dead of other causes), NED (no evidence of disease), and AWD (alive with disease) were coded as non-events. Deaths from other causes were censored at death for DSS. RFS used time to first recurrence or last documented recurrence-free follow-up. A missing or nonpositive recurrence-free interval was replaced with the OS interval only for patients without recurrence. Recurrence was the sole RFS event. Missing event indicators and missing or nonpositive times were excluded. A sensitivity analysis censored death without recurrence at the earlier of recorded recurrence-free follow-up and death.

Boundary distances, signal summaries, feature selection, and the intensity-adjusted comparisons were exploratory and were not preregistered

Univariate screening: We tested 50 exported boundary columns and 30 cisMI features per endpoint using univariate Cox regression.^23^ Continuous predictors were standardized using the population SD within the endpoint-specific analytic sample. Benjamini-Hochberg correction was applied jointly to the 80 tests within each endpoint at q < .05.^24^ Lead boundary features were selected by the lowest univariate P value. Reported effect estimates and P values for selected features are exploratory and do not account for selection.

Multivariable models: The base model used AJCC 8th-edition stage I-IV as one ordinal covariate, with IVA-IVC collapsed into IV. Stage-only and stage-plus-boundary models used the same complete-case patients within each endpoint. Continuous covariates were standardized on that sample, and Cox models used an L2 penalizer of 0.01. Separate models adjusted the lead boundary feature for stage and either mean signal in the same cumulative band or mean signal across the stromal compartment.

Internal validation: Apparent discrimination was summarized with Harrell’s c-index.^25–26^ Internal validation used 50 repeated 5-fold cross-validations (seed 42), stratified by event status. Within each training partition, all 50 boundary columns were re-screened by univariate Cox regression using model-based standard errors, and the lowest-P feature was selected. Standardization and both models were fitted in the training partition and evaluated on identical held-out patients. Fold concordance counts were pooled within each repeat. We also evaluated fixed 35-μm SD and p99 models on the same splits as exploratory comparisons; these features were not selected in an independent cohort. The 2.5th and 97.5th percentiles across repeats describe split variability and are not sampling confidence intervals. Cross-validation evaluates feature selection within this dataset and does not validate transportability to another institution.

Kaplan-Meier analysis: For visualization, patients were dichotomized at the median of each spatial feature. Survival curves were estimated by the Kaplan-Meier method,^27^ and groups were compared using the log-rank (Mantel-Cox) test. We did not optimize the dichotomization cutpoint; the median was chosen a priori.

Diagnostics and sensitivity analyses: We assessed proportional hazards using rank-transformed Schoenfeld residual tests^28^ for each covariate in the main models. Stage-stratified models included the lead boundary feature, an L2 penalizer of 0.01, and separate baseline hazards for stages I-IV. TMA-stratified models included the lead marker and a separate baseline hazard for each TMA, without a penalty; TMA number was used as a staining-batch proxy. We assessed correlations between the lead marker and mean signal using Spearman correlations.

Treatment-adjusted models included ordinal stage, the lead boundary feature, adjuvant radiation, and chemotherapy. Stage-interaction models included ordinal stage, an early-stage (I-II) indicator, the standardized feature, and the feature-by-early-stage interaction. Both model types used an L2 penalizer of 0.1; the interaction was assessed by its Wald test.

Missing-stage sensitivity used 20 Bayesian-ridge posterior-predictive imputations conditioned on the lead marker, event indicator, and Nelson-Aalen cumulative hazard.^29^ Predictors were standardized before fitting the imputation model to observed-stage records. Normal predictive draws were rounded and bounded to stages I-IV, using seed 42. Each completed dataset was analyzed with ordinal stage, the lead feature, and an L2 penalizer of 0.1. Outcomes and missing marker measurements were not imputed. Coefficients were pooled using Rubin’s rules. The two missing-stage records in each endpoint-specific marker sample were included in this sensitivity. Sample sizes: The eligible cohort comprised 133 patients, including 131 with boundary features. Lead-feature univariate models included 105 patients with 43 events for RFS, 119 with 43 events for OS, and 107 with 23 events for DSS. Matched stage-only and stage-plus-boundary models used 103 patients with 42 events, 117 with 43 events, and 105 with 23 events, respectively. The same patient sets contributed to the held-out comparisons. The number of events limited model complexity.

Software: Image analysis and cell segmentation were performed in QuPath^21^ v0.6.0. Spatial co-organization was computed with PySpace v0.1.0, a Python implementation of the SPACE algorithm.^22^ Boundary distance calculations used scipy.spatial^30^ (scipy 1.17.1). Survival analyses and statistical modeling were performed in Python 3.14.4 using lifelines^31^ 0.30.1 for Cox regression and Kaplan-Meier estimation, scipy^30^ for statistical tests, scikit-learn^32^ 1.8.0 for model evaluation metrics, and numpy^33^ 2.4.2 and pandas^34^ 2.3.3 for data processing. Figures were generated with matplotlib^35^ 3.10.8 and seaborn^36^ 0.13.2. All statistical tests were two-sided, with P < .05 considered statistically significant unless otherwise specified. The PySpace source code is publicly available at https://github.com/jcwal1516/PySpace.

## Results

### PySpace and the spatial analysis workflow

PySpace implements the SPACE algorithm in Python and is publicly available at https://github.com/jcwal1516/PySpace. We combined cisMI analysis in PySpace with a boundary-distance workflow that maps QuPath-exported cell coordinates and *SULF1* signal to annotated tumor-stroma interfaces (Figure 1). In the OSCC cohort, the boundary workflow generated distance profiles and cumulative-band summaries for survival analysis; cisMI analyses were exploratory.

**Figure 1.**
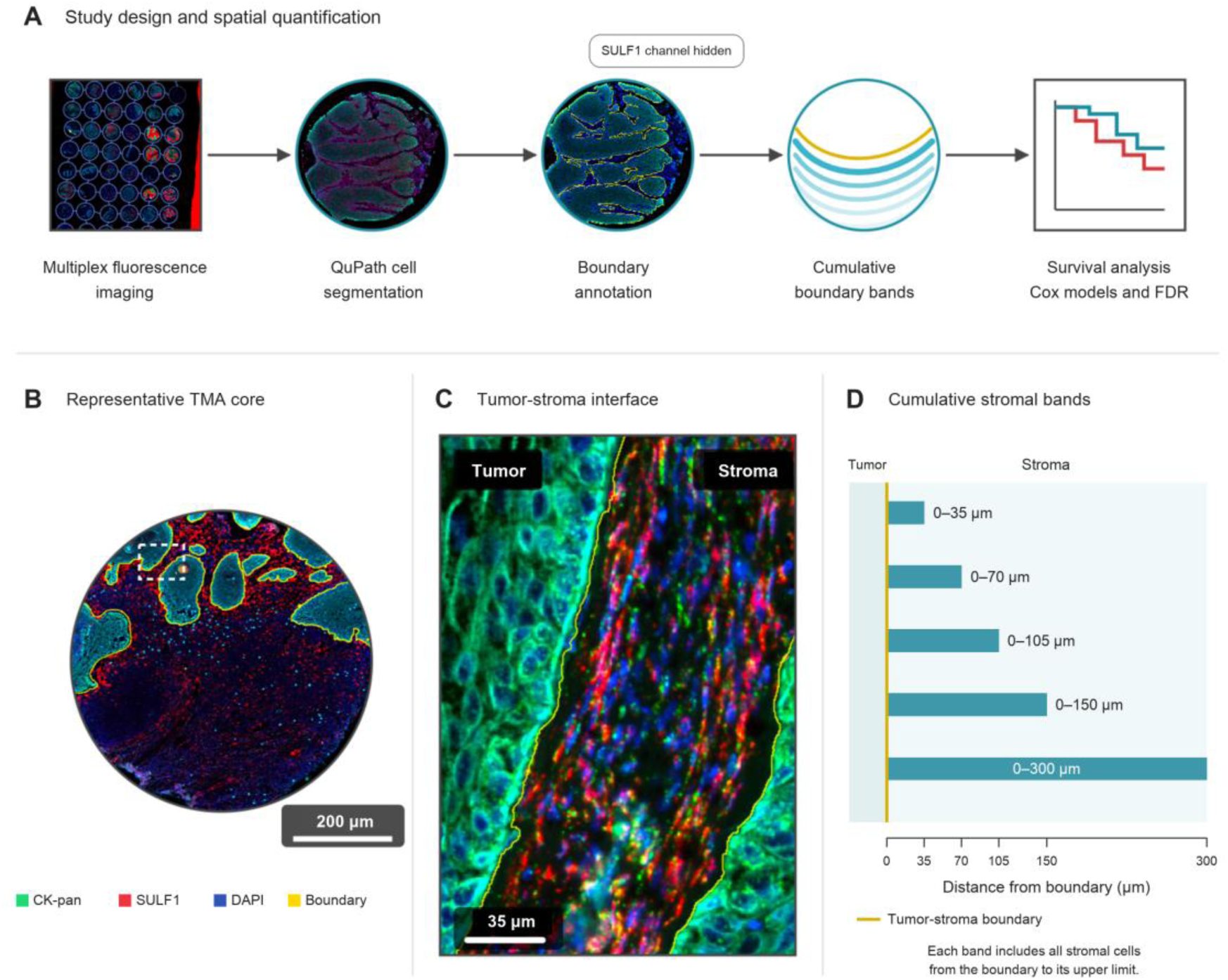
Study design and spatial quantification. (A) Primary-site TMA imaging, QuPath segmentation, blinded boundary annotation, cumulative boundary bands, and survival analysis. (B-C) Representative core and tumor-stroma interface. The red channel is RNAscope *SULF1* transcript signal; CK-pan marks tumor epithelium and DAPI marks nuclei. (D) Each horizontal bar represents a cumulative stromal band extending from the boundary to 35, 70, 105, 150, or 300 μm. The yellow line denotes the tumor-stroma boundary; bar lengths indicate distance, not cell density or signal intensity. Scale bars are shown in panels B and C.

### Cohort and spatial features

The eligible cohort included 133 patients and 218 primary-site cores after exclusion of non-SCC and in-situ/Stage 0-CIS records. Boundary measurements were available for 215 cores from 131 patients. There were 107 patients evaluable for recurrence, 121 for OS, and 109 for DSS before requiring marker availability. Median recorded OS follow-up was 3.53 years (range, 0.02-19.38). Table 1 summarizes the cohort.

**Table 1.**
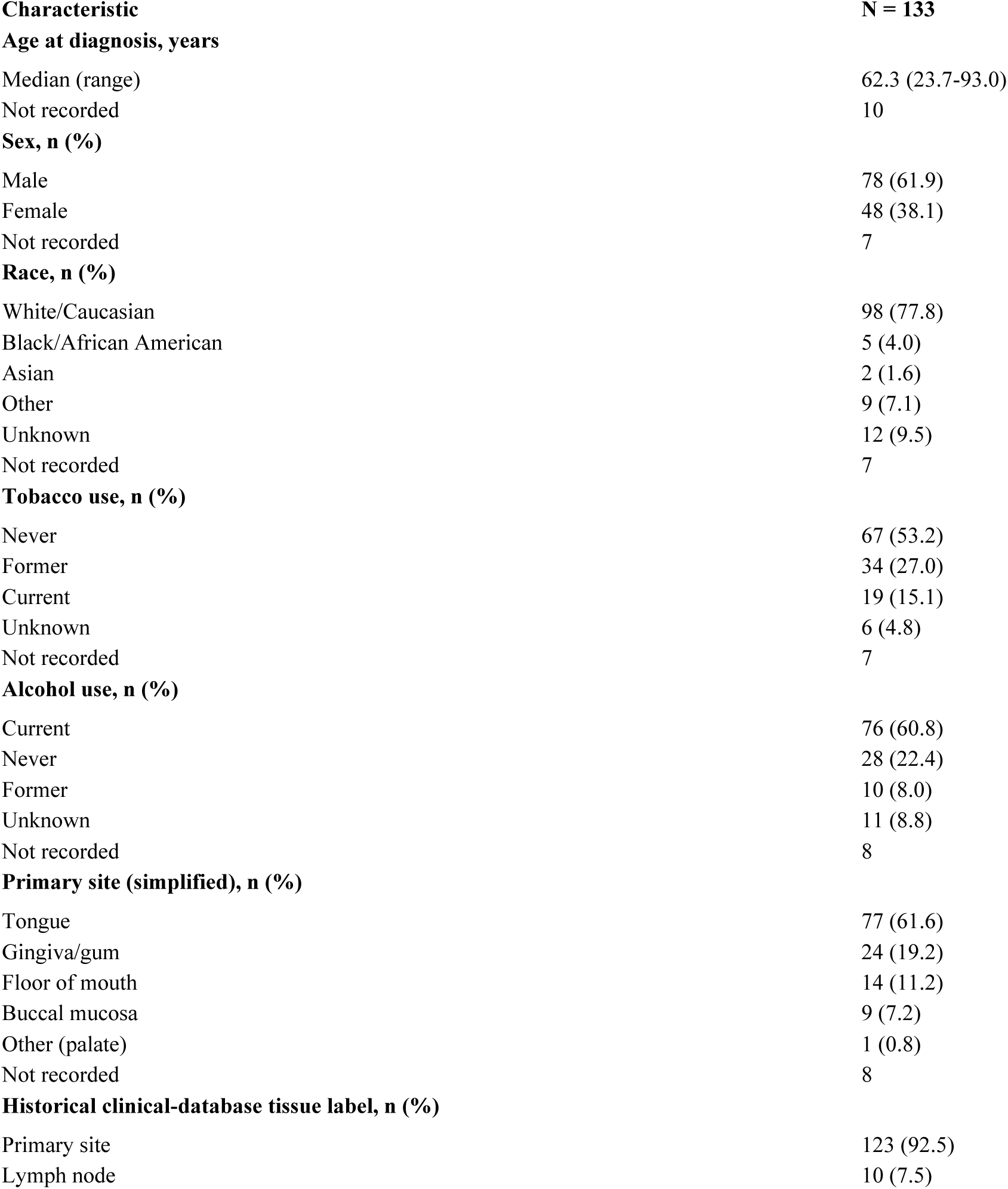
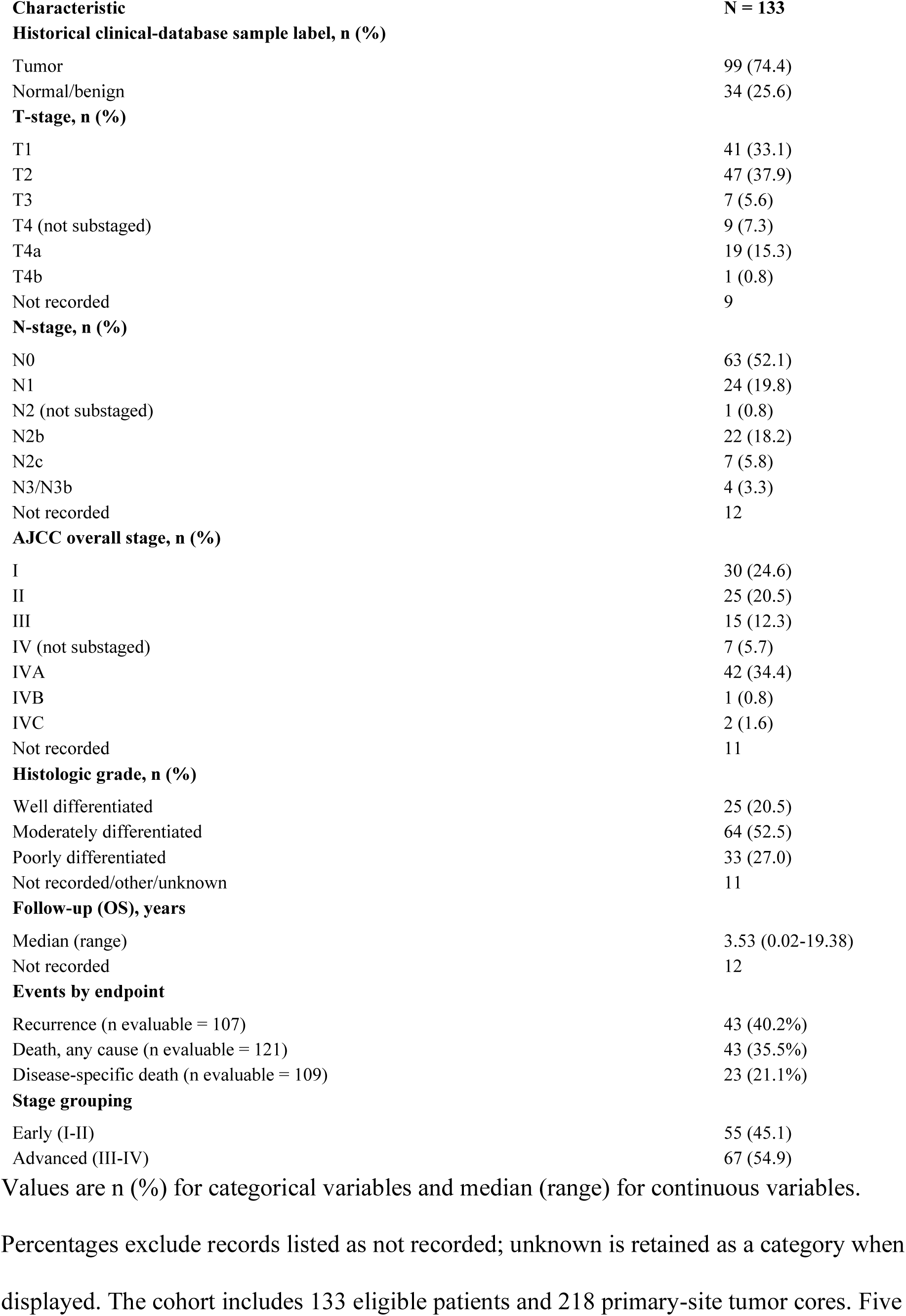
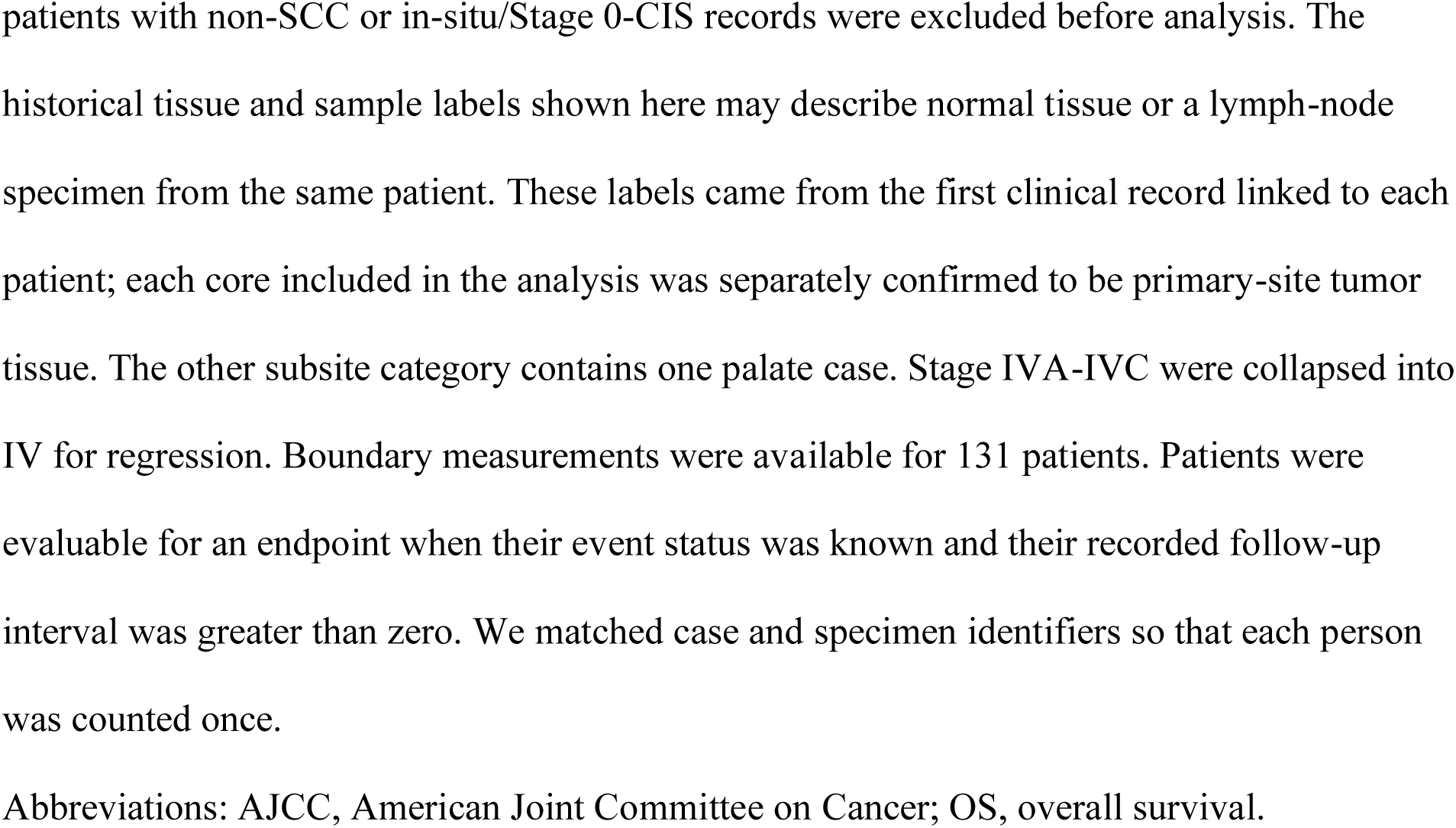
Clinicopathologic characteristics of the eligible primary-site cohort.

### Distribution of peritumoral *SULF1* transcript signal

In the 215 evaluable cores from 131 patients, the patient-averaged distance profile rose at the tumor-stroma interface and peaked in the 10-20 μm stromal bin (mean, 12.36 arbitrary units [a.u.]); mean signal was 9.70 a.u. at 100-110 μm and 7.87 a.u. at 290-300 μm (Figure 2C). The latter bin included 79 patients, compared with 131 at the peak. Within the cumulative 150-μm stromal band, the median patient-level signal-versus-distance slope was -0.0098 a.u./μm (IQR, - 0.0330 to 0.0029), indicating variation among patients in the direction and steepness of the gradient.

**Figure 2.**
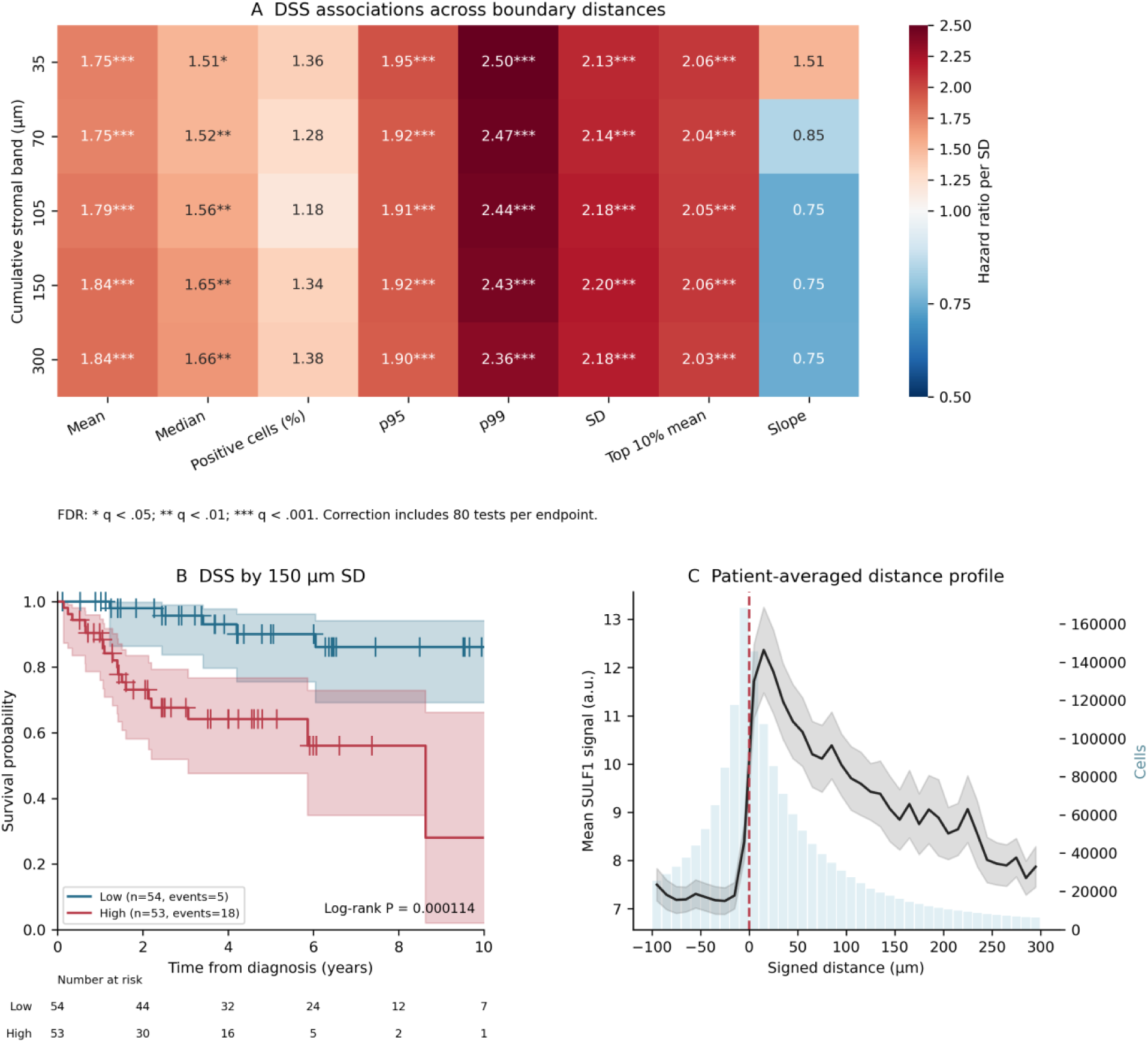
Boundary SULF1 transcript signal and disease-specific survival. (A) Univariate DSS hazard ratios per SD across cumulative stromal bands, with a color scale covering the complete observed range. Symbols denote FDR correction across 80 tests per endpoint. (B) DSS by median 150-μm SD, displayed through 10 years, with censor marks, 95% confidence bands, and numbers at risk aligned to the axis ticks. The log-rank test uses the full recorded follow-up. (C) Distance profiles from the 215 retained cores with exported measurements. Per-core means in 10-μm bins were averaged within patients, then across patients; shading is the SEM across contributing patients. Bars show pooled cell counts per bin without normalization to sampled tissue area. The 10-20 μm stromal bin includes 131 patients, and the 290-300 μm bin includes 79. Positive distance denotes stroma. SEM is descriptive and does not represent assay reproducibility.

Across the same 131 patients, median core-averaged mean signal was 8.14 a.u. in stroma (IQR, 6.60-12.28) and 6.75 a.u. in tumor (IQR, 5.80-8.09). The median within-patient stromal-to-tumor ratio was 1.27 (IQR, 1.02-1.63). Tumor-side signal was lower on average, but these measurements do not establish absence of transcript in tumor cells. Within 150 μm, the median patient-level GMM-defined positive-cell fraction was 14.7% (IQR, 11.3-20.5); this relative threshold does not define absolute transcript positivity. Figure 2C shows cell counts per bin, not cell density per tissue area.

### Boundary transcript signal and survival

The screen evaluated 50 boundary columns and 30 cisMI features per endpoint. For RFS, the lead feature was 70-μm SD (n = 105; 43 events; HR = 1.70 per SD; 95% CI, 1.34-2.16; q = 2.5 × 10⁻⁴). For OS, the lead feature was 150-μm SD (n = 119; 43 events; HR = 1.87 per SD; 95% CI, 1.49-2.37; q = 4.1 × 10⁻⁶). For DSS, the lead feature was 150-μm SD (n = 107; 23 events; HR = 2.20 per SD; 95% CI, 1.64-2.94; q = 2.1 × 10⁻⁶). Figure 2 shows the DSS screen, survival curves, and patient-averaged distance profile. Correlation across cumulative bands limits interpretation of any one distance as a distinct biological threshold.

### Comparison with mean signal

In exploratory DSS models adjusted for stage and mean signal, the 150-μm SD association remained with either same-band mean signal (HR = 4.04; 95% CI, 1.65-9.91; P = .0022) or overall stromal mean signal (HR = 3.91; 95% CI, 1.65-9.25; P = .0019). The correlations with these means were high (Spearman ρ = .893 and .870), and the conditional estimates were imprecise.

### Internal validation

Adding the lead feature increased apparent discrimination in the matched stage models (Table 2). In the stage-adjusted DSS model, 150-μm SD remained associated with outcome (HR = 2.11; 95% CI, 1.55-2.86). For RFS, mean held-out c-index increased from 0.553 to 0.626 (ΔC = +0.072; repeat percentiles, +0.004 to +0.119). For OS, mean held-out c-index increased from 0.606 to 0.673 (ΔC = +0.067; repeat percentiles, +0.026 to +0.101). For DSS, mean held-out c-index increased from 0.613 to 0.754 (ΔC = +0.141; repeat percentiles, +0.105 to +0.192). Fixed 35-μm SD and p99 models gave DSS ΔC values of +0.147 and +0.144. The 150-μm SD feature was selected in 39.2% and 150-μm p99 in 28.0% of DSS training folds; no other feature exceeded 10% (Figure 3B). These results support internal consistency across correlated features, with uncertainty about the preferred distance and summary statistic.

**Figure 3.**
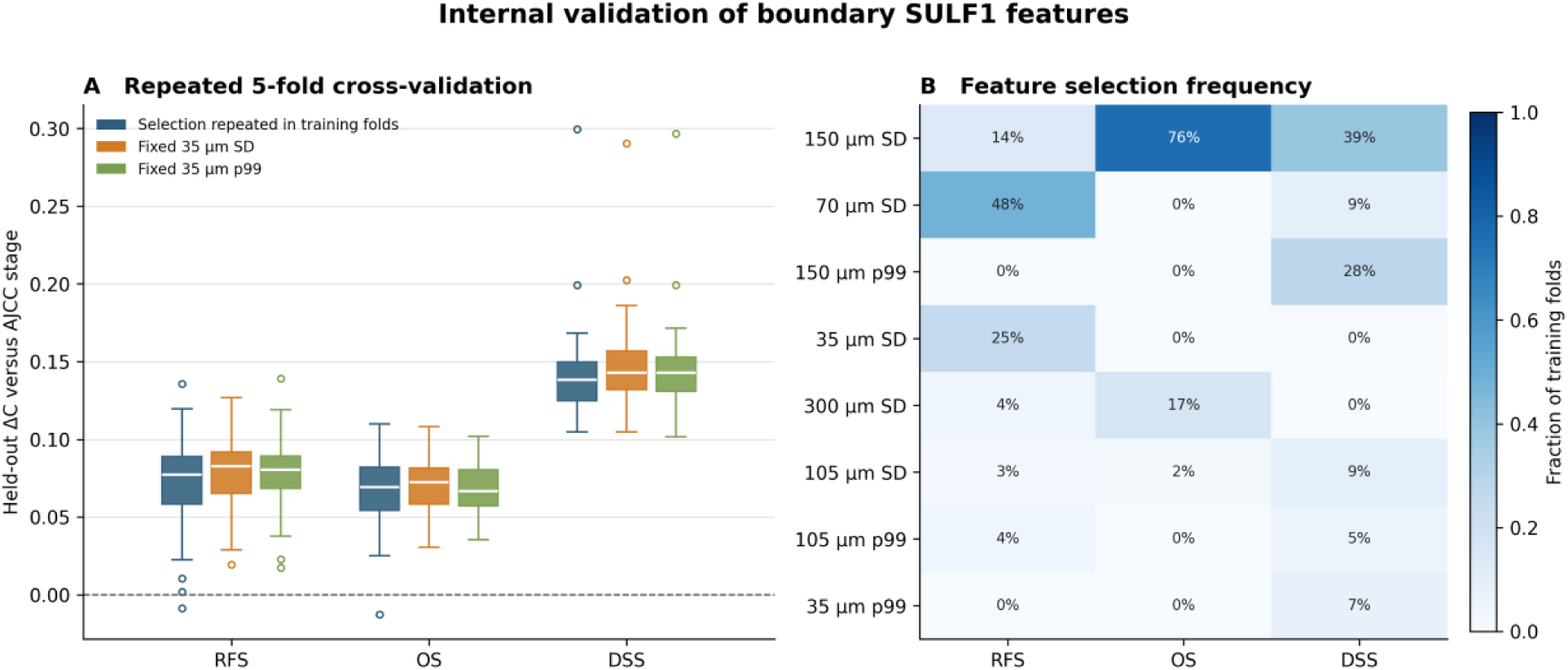
Internal validation of boundary *SULF1* features. (A) Held-out ΔC distributions across 50 repeated 5-fold cross-validations, comparing training-fold-selected boundary features with fixed 35-μm SD and p99 models. Boxes show the interquartile range and median; whiskers extend to the most extreme values within 1.5 interquartile ranges, and outliers are shown as open circles. (B) Selection frequencies for the eight features with the highest mean frequency across endpoints. Omitted features account for any remaining percentage. Each endpoint includes 250 training folds. Percentile ranges in the text and Table 2 describe variation across repeats and are not confidence intervals.

**Table 2.** AJCC-stage-based discrimination with boundary *SULF1* features.

| Endpt. | N / events | Lead boundary feature | Apparent C: stage / augmented | Held-out C: stage / augmented | Held-out $\Delta C$ (repeat percentiles) |
| --- | --- | --- | --- | --- | --- |
| RFS | 103 / 42 | 70-μm SD | 0.556 / 0.656 | 0.553 / 0.626 | +0.072 (+0.004 to +0.119) |
| OS | 117 / 43 | 150-μm SD | 0.607 / 0.690 | 0.606 / 0.673 | +0.067 (+0.026 to +0.101) |
| DSS | 105 / 23 | 150-μm SD | 0.617 / 0.764 | 0.613 / 0.754 | +0.141 (+0.105 to +0.192) |

Stage-only and stage-plus-boundary models used identical patients within each endpoint.
Apparent c-indices use the full-sample-selected feature named in the table. Held-out c-indices are means across 50 repeated 5-fold cross-validations, with all 50 boundary columns re-screened within each training fold; the selected feature could therefore differ from the full-sample lead.
Parentheses contain the 2.5th and 97.5th percentiles across repeats, which describe split variability and are not confidence intervals. All models used ordinal AJCC stage I-IV and an L2 penalizer of 0.01.

### Sensitivity analyses

In patients with recorded adjuvant radiation and chemotherapy status, the lead boundary HRs after adjustment for stage and both treatments were 1.64 (95% CI, 1.27-2.11) for RFS (n = 102; 42 events), 1.90 (95% CI, 1.47-2.47) for OS (n = 106; 34 events), and 1.93 (95% CI, 1.43-2.60) for DSS (n = 103; 23 events). With the two missing-stage values imputed in each endpoint-specific sample, the HRs were 1.54 (95% CI, 1.22-1.95) for RFS, 1.65 (95% CI, 1.31-2.08) for OS, and 1.84 (95% CI, 1.38-2.44) for DSS.

Feature-by-stage interaction tests were not significant (RFS P = .644; OS P = .837; DSS P = .765). AJCC stage violated proportional hazards in the DSS stage-only model (P = .027); the boundary feature did not (P = .595 in the augmented model). The DSS boundary association remained after stratifying baseline hazards by stage (HR = 1.86; 95% CI, 1.37-2.52). With TMA-stratified baseline hazards, the RFS estimate was borderline (HR = 1.39; 95% CI, 1.00-1.94; P = .050), whereas OS (HR = 1.89; 95% CI, 1.36-2.63; P < .001) and DSS (HR = 2.16; 95% CI, 1.41-3.31; P < .001) remained associated.

Censoring death without recurrence at the earlier of recorded recurrence-free follow-up and death yielded results identical to the primary RFS analysis because no censoring times changed (70-μm SD: HR = 1.70 per SD; 95% CI, 1.34-2.16; n = 105; 43 events).

### Exploratory cisMI associations

The 35-μm tumor-with-tumor-linked cisMI feature had the lowest univariate P value among the 30 cisMI candidates for all three endpoints. It was associated with RFS (HR = 0.51 per SD; 95% CI, 0.36-0.72; n = 105; 43 events; q = 5.2 × 10⁻⁴); OS (HR = 0.59 per SD; 95% CI, 0.43-0.82; n = 119; 43 events; q = 3.6 × 10⁻³); and DSS (HR = 0.35 per SD; 95% CI, 0.21-0.59; n = 107; 23 events; q = 2.4 × 10⁻⁴). These were same-cohort exploratory associations; cisMI was not evaluated in the boundary-feature cross-validation.

## Discussion

Peritumoral *SULF1* signal dispersion was associated with recurrence, overall mortality, and disease-specific death in this retrospective cohort. The largest internally validated discrimination gain was for DSS. The 150-μm SD feature remained associated with DSS after stage adjustment (HR = 2.11; 95% CI, 1.55-2.86). Multiple correlated SD and upper-tail measurements carried similar information, and the preferred feature varied across training folds.

Greater spatial co-organization of tumor-cell proportion and tumor-associated *SULF1* signal, measured by cisMI, was associated with better outcomes across all three endpoints. The cisMI associations were exploratory and were not cross-validated; the prediction analyses therefore focused on boundary features, which underwent repeated cross-validation. Coverage varied by radius, limiting comparisons across spatial scales.

This study extends the 20-specimen Yang et al. pilot, which localized *SULF1* transcript to stromal cells by RNAscope,^8^ to a 133-patient eligible cohort. The present assay also measures RNA in situ; it does not quantify SULF-1 protein. Mechanistic studies provide context: *SULF1* knockout in CAFs reduced invasion in HNSCC models.^15^ The colorectal study further supports a CAF-associated SULF-1 program involving extracellular-matrix remodeling and angiogenesis.^16^ Together, these observations support, but do not prove, the interpretation that boundary *SULF1* transcript signal marks a host stromal program. These data support further mechanistic evaluation of stromal SULF signaling as an experimental target.

The distance profile places the highest average *SULF1* transcript signal immediately outside the tumor boundary, while the survival-associated SD measures cell-to-cell variation within that stromal region. SD remained associated with DSS after adjustment for same-band or overall stromal mean signal, suggesting that average intensity alone does not fully explain the association. However, SD depends on signal scale, and its strong correlation with mean signal limits separation of their effects. The gain in held-out discrimination was measured against AJCC stage alone; superiority to a stage-plus-mean-signal model was not tested in cross-validation. The data therefore do not establish that boundary sampling is superior to other stromal regions. A direct comparison with SULF-1 protein immunohistochemistry would also require a separate assay, because the present study measures RNA.

Multiparameter imaging has linked integrated tumor-stromal phenotypes to HNSCC recurrence and outcome.^37^ Imaging mass cytometry in OSCC has also identified an invasive-margin lymphatic endothelial marker associated with outcome in an independent cohort.^38^ Our study examines *SULF1* transcript signal within distance-defined stromal bands. Whether this measurement adds information to these broader cellular signatures remains unknown.

This is a single-institution retrospective TMA study without external validation, with only 23 disease-specific deaths. The cohort spans specimens accessioned over 25 years, and clinical follow-up durations vary. Repeated cross-validation accounts for selection within this dataset; its correlated repeats do not provide sampling confidence intervals or establish external reproducibility. TMAs incompletely sample intratumoral heterogeneity, cumulative bands overlap, and p99 estimates from small cell counts may be unstable. The six TMAs were stained in separate runs, and no post hoc per-TMA signal normalization was applied. Boundaries were drawn by one investigator and reviewed by a second; independent re-annotation and numeric agreement were unavailable. The stromal compartment was defined geometrically and was not restricted by a CAF-specific marker. The ordinal stage comparator does not represent a complete clinicopathologic prediction model. Intensity-adjusted models have limited precision, and the missing-stage imputation uses an approximate model for an ordinal variable.

Peritumoral *SULF1* signal dispersion was associated with OSCC outcomes, with the largest internal discrimination gain for DSS. The findings justify evaluation of a fixed assay and analysis specification in an independent cohort before clinical use.

## Declarations

### Ethics approval and consent to participate

This study was approved by the Georgetown-MedStar institutional review board (IRB 1992-48). Informed consent was obtained from all participants. The study was conducted in accordance with the Declaration of Helsinki.

### Author contributions

Gregory Furletti: Formal analysis, Methodology, Visualization, Writing – original draft. Trystan Conrad Guerrero: Formal analysis, Validation, Blinded review of the tumor-stroma annotations, Writing – review & editing. Edward C. Schrom: Software, Writing – review & editing. Radoslav Goldman: Conceptualization, Funding acquisition, Supervision, Writing – review & editing.

Bruce Davidson: Conceptualization, Writing – review & editing. Bhaskar Kallakury: Investigation, Resources, Writing – review & editing. All authors reviewed and approved the final manuscript.

## Acknowledgments

We thank Aaron M. Rozeboom and the Histopathology and Tissue Shared Resource at the Georgetown University Lombardi Comprehensive Cancer Center for assistance with data acquisition.

## Data availability

The PySpace source code is publicly available at https://github.com/jcwal1516/PySpace. Raw imaging and patient-level study data are not publicly posted and are available from the corresponding author upon reasonable request and institutional approval.

Declaration of generative AI and AI-assisted technologies in the writing process

During preparation of this manuscript, the authors used OpenEvidence for literature review and source finding. After using these tools, the authors reviewed and edited the content as needed and take full responsibility for the content of the publication.

